# Lifestyle patterns in the Peruvian population: analysis of tobacco, alcohol and fruit/vegetable consumption based on a nine-year national survey

**DOI:** 10.1101/2024.05.26.24307960

**Authors:** Víctor Juan Vera-Ponce, Fiorella E. Zuzunaga-Montoya, Luisa Erika Milagros Vásquez-Romero, Joan A. Loayza-Castro, Carmen Inés Gutierrez De Carrillo, Stella M. Chenet

## Abstract

**Introduction:** A population’s lifestyle is a factor in public health and general well-being; thus, three indicators are alcohol consumption, smoking activity, and a healthy diet.

**Objectives:** The study aimed to determine the trends and factors associated with alcohol consumption, smoking activity, and fruit and vegetable consumption.

**Methods:** This cross-sectional study was based on the National Survey of Demography and Family Health (ENDES) database between 2014 and 2022. Smoking status was categorized as never smoker, former smoker, current smoker, and daily smoker; alcohol consumption was classified as not having consumed in the last 12 months, non-excessive consumption in the previous 30 days, and excessive consumption. Fruit and vegetable consumption was divided into whether five or more servings were consumed, and it was formed separately into three or more portions for fruit and two more portions for vegetables.

**Results:** The overall prevalence of lifestyles, starting with daily smoking, excessive alcohol consumption, and consumption of fewer than five portions of fruits and vegetables per day, are 1.55%, 3.06%, and 91.02%, respectively. Significant differences were found between each of them in terms of age, sex, and socioeconomic status.

**Conclusions:** The current study observed slight changes in the prevalence of daily and current smokers. Additionally, an increase in excessive alcohol consumption in 2020, followed by a decrease in 2022, was found. The study draws attention to the troubling scarcity of fruit and vegetable intake within the population, underscoring potential health risks posed by inadequate consumption of these nutritious foods. The study not only found significant divergences in wellness practices relating to gender, age, and area but also highlighted how accounting for such elements in public health intervention strategies is paramount by emphasizing the need to consider those variances.

## Introduction

A population’s lifestyle is a determining factor in public health and general well-being. Thus, three indicators related to alcohol consumption, smoking activity, and consumption of healthy foods, such as fruits and vegetables ^(1)^.

Alcohol consumption use throughout society marks it as a meaningful public health measure, with its influences on communities having been thoroughly recorded. Globally, the widespread consumption of alcohol results in a significant amount of preventable illness and loss of life well before what would otherwise be considered normal. The World Health Organization (WHO) indicates that alcohol consumption is linked to over 200 health conditions and represents a primary risk element for various non-communicable diseases, specifically cardiovascular diseases, cancers, and liver disorders ^(2)^. While widespread smoking continues posing grave risks to individuals and the public alike, efforts to curb this preventable health crisis remain as urgent as ever. The WHO ^(3)^ has reported that the leading preventable cause of death worldwide continues to be smoking, as it plays a significant role in significantly increasing the risk of developing respiratory illnesses, heart conditions, and various forms of cancer - all of which could otherwise often be avoided.

Studies into fruit and vegetable consumption have conclusively demonstrated their significant role in forestalling chronic illnesses and cultivating robust well-being, with a plethora of examinations uniformly underlining the vital importance of such nutritious items. Regularly consuming sufficient fruits and vegetables has decreased the likelihood of developing certain health conditions, such as cardiovascular diseases ^(4)^.

This study utilizes data collected by ENDES between 2014 and 2022 to analyze these three critical aspects of lifestyle in the Peruvian population, which are crucial elements in determining health risks and promoting healthy habits. Analyzing current health trends in Peru more closely aims to furnish a more nuanced view that yields meaningful perspectives on crafting public health initiatives and interventions better tailored to the nation’s needs. By concentrating on a particular group, this study provides a more nuanced perspective on how specific risk factors and health behaviors may manifest differently when viewed through a contextual lens. Understanding these patterns is essential for formulating targeted and effective public health interventions, especially in a country with as rich cultural and geographical diversity as Peru.

## Material and Methods

### Design

We conducted an analytical cross-sectional study using the National Survey of Demography and Family Health (ENDES) database between 2014 and 2022 ^(5)^. Our study adhered to the STROBE guidelines to ensure a comprehensive reporting of observational studies in epidemiology ^(6)^.

### Population, Eligibility Criteria, and Sample

The survey was conducted nationally and included Peruvians aged 15 to 49 from urban and rural areas across 24 departments in Peru. ENDES employed a two-stage stratified sampling method, independently probabilistic for rural and urban areas ^(5)^. Individuals of all ages were included, while those lacking the main variables of interest were excluded.

### Assessment of consistency and plausibility of measurements

We established parameters to ensure that only practical blood pressure readings were included, following methodologies from related previous analyses. Systolic blood pressure (SBP) measurements were required to be between 70 and 270 mmHg, and diastolic blood pressure (DBP) was between 50 and 150 mmHg. Measurements not meeting these criteria were excluded from the analysis ^(7)^.

### Variables and measurement

The main variables were smoking status, alcohol consumption, and fruit/vegetable consumption.

- Smoking Status: Obtained from respondent self-report, categorized as never smoked, ex-smoker, current smoker, and daily smoker ^(8)^. For regression analysis, this was dichotomized into never/ex-smoker versus current/daily smoker.
- Alcohol Consumption: Defined by self-report, categorized into never or not used in the last 12 months; non-excessive use (≥ one occasion in the previous 30 days, but <5 drinks in men or <4 in women in the last 12 months) and excessive use (≥ one occasion in the previous 30 days and ≥ five drinks in men or ≥ four drinks in women). For regression analysis, this was dichotomized by combining the first two versus excessive consumption ^(9)^.
- Fruit and Vegetable Consumption: Classically worked with the consumption of ≥ five portions per day (yes versus no), and separately with the consumption of fruits ≥ 3 portions per day and vegetables ≥ 2 portions per day.

Covariables such as sex, age group, marital status, region, wealth index, educational level, urban or rural residence, physical disability, presence of type 2 diabetes, presence of arterial hypertension, altitude ranges, race, and nutritional status were considered.

Nutritional status, evaluated through Body Mass Index (BMI), with cutoff points established by the WHO: average weight (< 25 kg/m²), overweight (BMI between 25 and 29.9 kg/m²), and obesity (BMI ≥ 30 kg/m²). Abdominal obesity was measured according to the Cholesterol Education Program Adult Treatment Panel III criteria (WC-ATPIII): waist circumference ≥ 102 cm in men and ≥ 88 cm in women ^(10)^.

### Statistical analysis

Version 4.03 of R software will perform various statistical analyses in this study. Firstly, a descriptive analysis of all variables will be carried out, providing an overview of the dataset’s characteristics. Concurrently, a trend chart for each primary variable was created.

Subsequently, a bivariate analysis of all the main variables will be conducted. Following this, a Poisson regression model with robust variance was implemented to delve deeper into the analysis of factors associated with each of the variables above. This model allowed for calculating adjusted Prevalence Ratios (aPRs) along with their 95% confidence intervals.

As part of the analysis of interrelations among the studied lifestyle factors, Venn diagrams were created. These diagrams were crucial for visualizing and understanding the intersections and relationships between smoking status, alcohol consumption, and fruit and vegetable consumption (using the dichotomized versions of the main variables). Additionally, a specific Venn diagram was created to examine the relationship between fruit and vegetable consumption, providing insights into the joint consumption patterns of these foods.

### Ethical considerations

The data used are public and anonymized, eliminating any ethical risk to participants. ENDES ensures informed consent from participants for collecting information through the survey. The database can be accessed through the following link: https://proyectos.inei.gob.pe/microdatos/

## Results

### Participants

A total of 309,652 participants were included in the study. The gender distribution was nearly equal, with 51.65% being women. Of these, 16.58% are older adults. Only 13.28% reside in the jungle regions. Regarding concomitant pathologies, 23.28% and 44.32% have obesity according to BMI and WC, respectively. The overall prevalence of lifestyles, starting with daily smoking, excessive alcohol consumption, and consumption of fewer than five portions per day of fruits and vegetables, are 1.55%, 3.06%, and 91.02%, respectively. Additionally, only 21.18% and 7.27% consume adequate portions of fruits and vegetables, respectively. The total characteristics can be seen in Table 1.

**Table 01.**
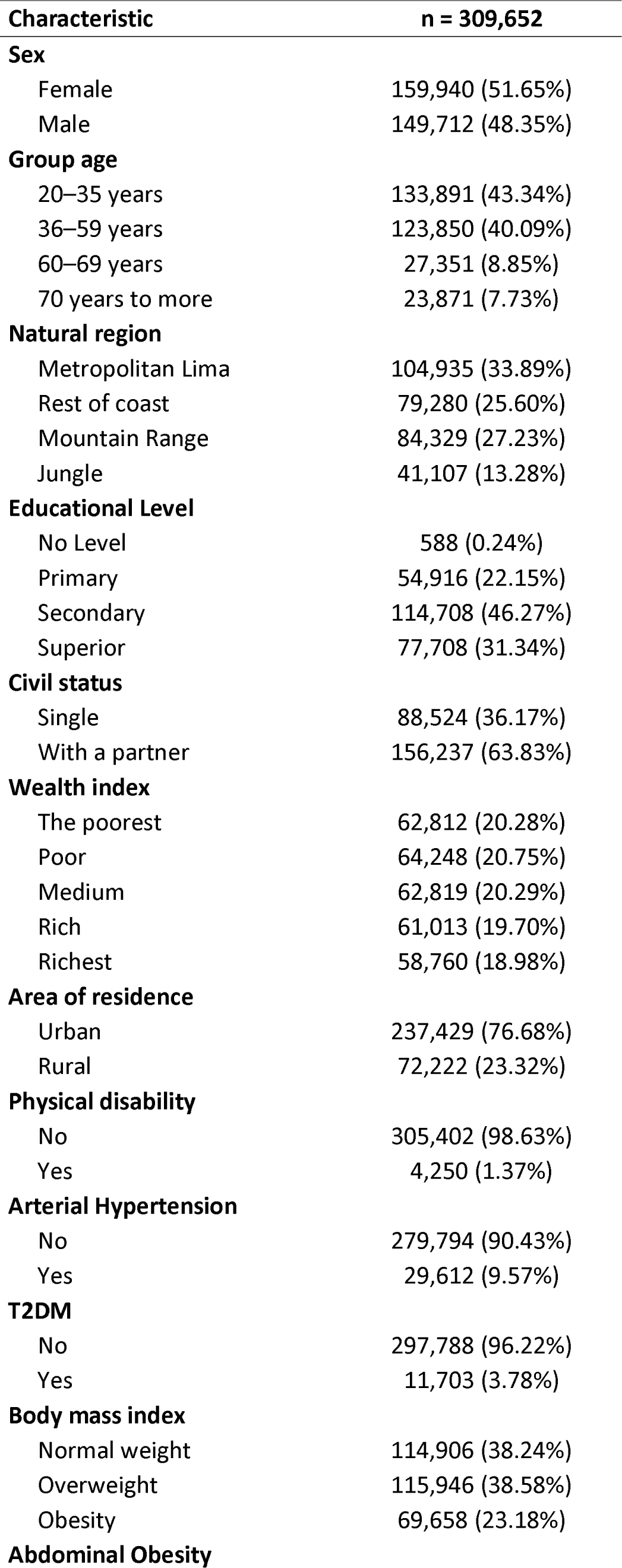
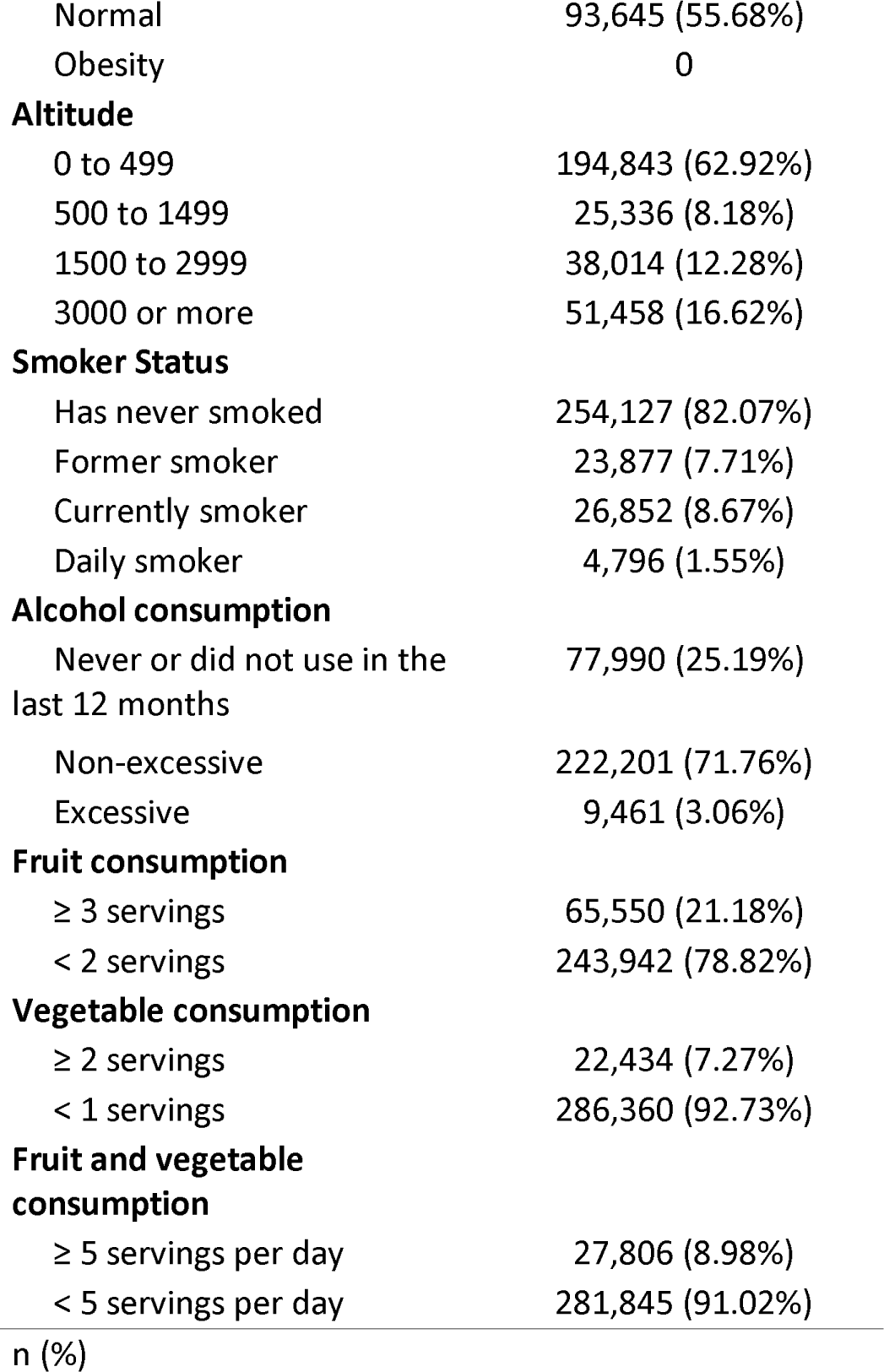
Descriptive characteristics of the study sample.

| Characteristic | n = 309,652 |
| --- | --- |
| **Sex** |  |
| Female | 159,940 (51.65%) |
| Male | 149,712 (48.35%) |
| **Group age** |  |
| 20–35 years | 133,891 (43.34%) |
| 36–59 years | 123,850 (40.09%) |
| 60–69 years | 27,351 (8.85%) |
| 70 years to more | 23,871 (7.73%) |
| **Natural region** |  |
| Metropolitan Lima | 104,935 (33.89%) |
| Rest of coast | 79,280 (25.60%) |
| Mountain Range | 84,329 (27.23%) |
| Jungle | 41,107 (13.28%) |
| **Educational Level** |  |
| No Level | 588 (0.24%) |
| Primary | 54,916 (22.15%) |
| Secondary | 114,708 (46.27%) |
| Superior | 77,708 (31.34%) |
| **Civil status** |  |
| Single | 88,524 (36.17%) |
| With a partner | 156,237 (63.83%) |
| **Wealth index** |  |
| The poorest | 62,812 (20.28%) |
| Poor | 64,248 (20.75%) |
| Medium | 62,819 (20.29%) |
| Rich | 61,013 (19.70%) |
| Richest | 58,760 (18.98%) |
| **Area of residence** |  |
| Urban | 237,429 (76.68%) |
| Rural | 72,222 (23.32%) |
| **Physical disability** |  |
| No | 305,402 (98.63%) |
| Yes | 4,250 (1.37%) |
| **Arterial Hypertension** |  |
| No | 279,794 (90.43%) |
| Yes | 29,612 (9.57%) |
| **T2DM** |  |
| No | 297,788 (96.22%) |
| Yes | 11,703 (3.78%) |
| **Body mass index** |  |
| Normal weight | 114,906 (38.24%) |
| Overweight | 115,946 (38.58%) |
| Obesity | 69,658 (23.18%) |
| **Abdominal Obesity** |  |
| Normal | 93,645 (55.68%) |
| Obesity | 0 |
| **Altitude** |  |
| 0 to 499 | 194,843 (62.92%) |
| 500 to 1499 | 25,336 (8.18%) |
| 1500 to 2999 | 38,014 (12.28%) |
| 3000 or more | 51,458 (16.62%) |
| **Smoker Status** |  |
| Has never smoked | 254,127 (82.07%) |
| Former smoker | 23,877 (7.71%) |
| Currently smoker | 26,852 (8.67%) |
| Daily smoker | 4,796 (1.55%) |
| **Alcohol consumption** |  |
| Never or did not use in the last 12 months | 77,990 (25.19%) |
| Non-excessive | 222,201 (71.76%) |
| Excessive | 9,461 (3.06%) |
| **Fruit consumption** |  |
| ≥ 3 servings | 65,550 (21.18%) |
| < 2 servings | 243,942 (78.82%) |
| **Vegetable consumption** |  |
| ≥ 2 servings | 22,434 (7.27%) |
| < 1 servings | 286,360 (92.73%) |
| **Fruit and vegetable consumption** |  |
| ≥ 5 servings per day | 27,806 (8.98%) |
| < 5 servings per day | 281,845 (91.02%) |

### Lifestyle Trends

In the case of smoking status, there is a progressive decrease in the percentage of people identifying as current smokers, with a significant reduction in the prevalence of current smokers in 2020, dropping to 6.28%, the lowest point in the entire time interval studied; however, this trend slightly reversed in 2021, with an increase in the proportion of current smokers to 6.70%, although this remains below the initial level of 2014. Similarly, the proportion of daily smokers also decreased, from 2.00% in 2014 to 1.37% in 2022. In contrast, the category of ex-smokers has shown minor fluctuations during this period, with an initial slight increase followed by a decrease and then another increase in 2022 to 8.13%—figure 1.

**Figure 1.**
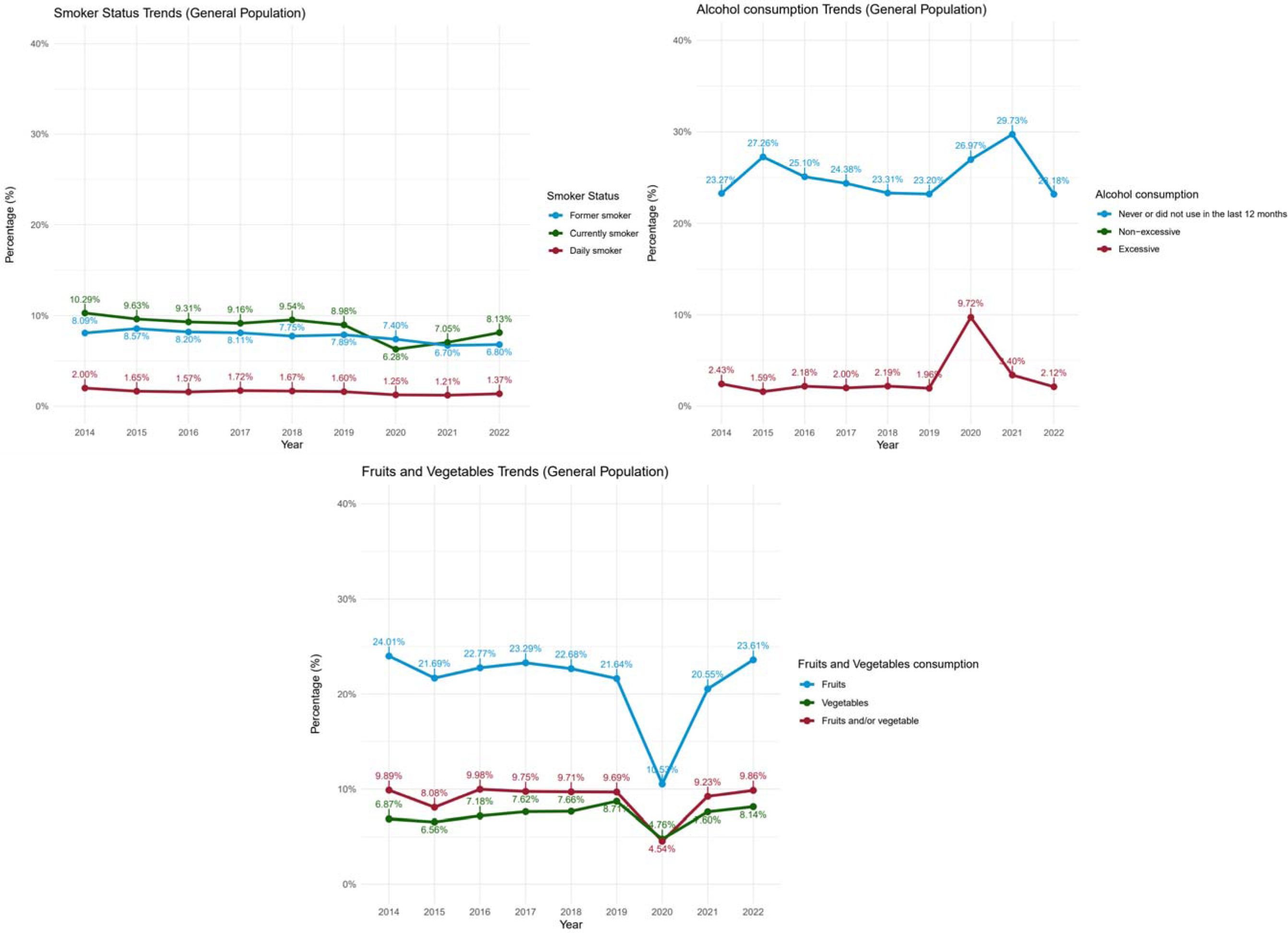
Trend graph of lifestyles in the Peruvian population

In the case of alcohol consumers, over the studied period, the proportion of individuals who have never consumed alcohol or did not in the last year shows moderate variability, with a decrease in 2022 to 23.20%. Notably, there was a peak in excessive consumption in 2020, which rose to 9.72%, followed by a significant decline to 2.12% in 2022, suggesting a marked reduction in extreme alcohol consumption behaviors after that year. Non-excessive consumption remained relatively stable, with a slight decrease over the years—figure 2.

**Figure 2.**
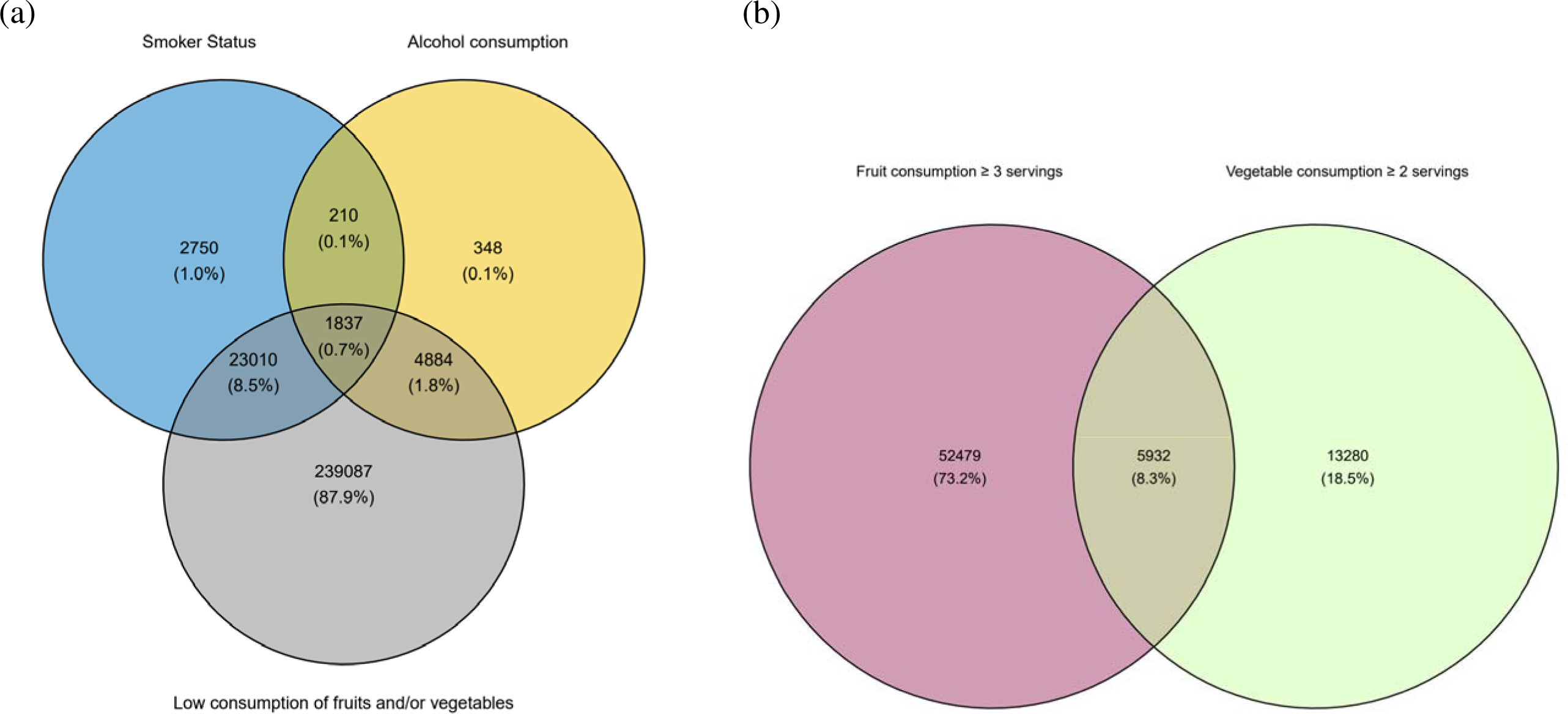
Venn diagram of (a) lifestyles and (b) consumption of fruits and vegetables

The general population’s fruit and vegetable consumption trend from 2014 to 2022 shows an interesting dynamic. Fruit consumption remained relatively stable, slightly decreasing in 2020 to 20.55%, but rebounded in 2022 to 23.61%. On the other hand, vegetable consumption experienced a notable fall in 2020, reaching its minimum at 4.54% and then slightly recovering to 8.14% in 2022. The category combining fruit and vegetable consumption also reflects this decrease in 2020 with 9.23%, followed by an increase in 2022 to 9.86%—figure 3.

### Relationship Between Lifestyles

The Venn diagram in Figure 4 illustrates the intersection between current or daily smokers, excessive alcohol consumers, and those with low fruit and vegetable consumption (defined as fewer than five portions daily). Of the total population analyzed, 87.9% do not identify with any of the mentioned risk behaviors. 8.5% of individuals are current or daily smokers and also have low fruit and vegetable consumption but do not consume alcohol excessively. 1.8% of participants drink alcohol excessively and do not meet the recommendation for fruit and vegetable consumption, though they are not current or daily smokers. Only 0.1% of the population combines the three risk behaviors: they are present or daily smokers, consume alcohol excessively, and have low fruit and vegetable consumption.

The Venn diagram in Figure 5 reflects how subjects who consume at least three portions of fruits and fewer than two portions of vegetables overlap. Generally, an overlap of 8.3% indicates that this percentage of the population meets both criteria.

### Factors Associated with Lifestyles

Table 2 presents the adjusted Poisson regression analysis results, examining the association between various sociodemographic factors and lifestyles. Additionally, bivariate analyses performed with the chi-squared test with Rao & Scott’s second-order correction are found in supplementary materials 1 and 2.

**Table 02.**
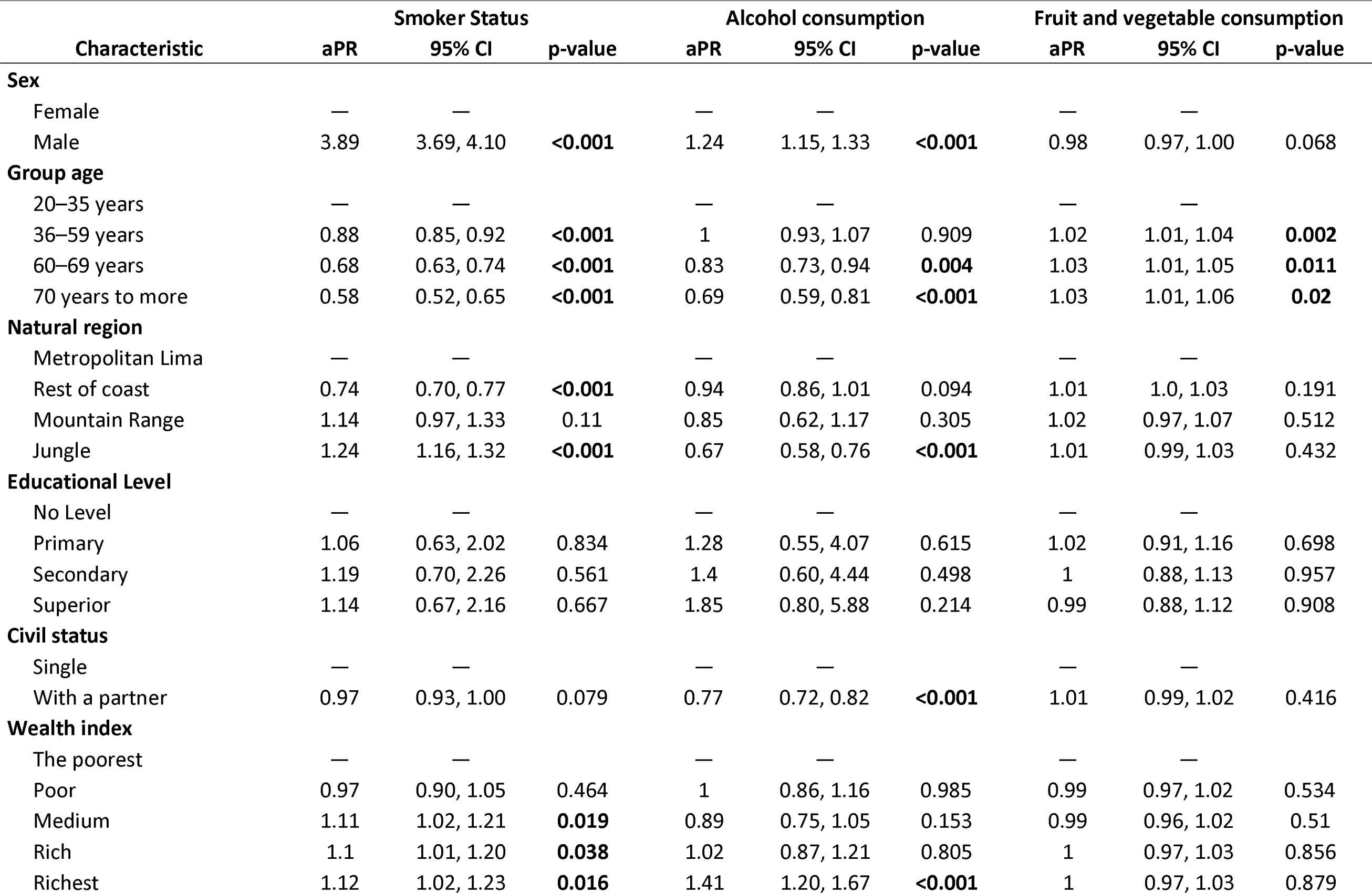
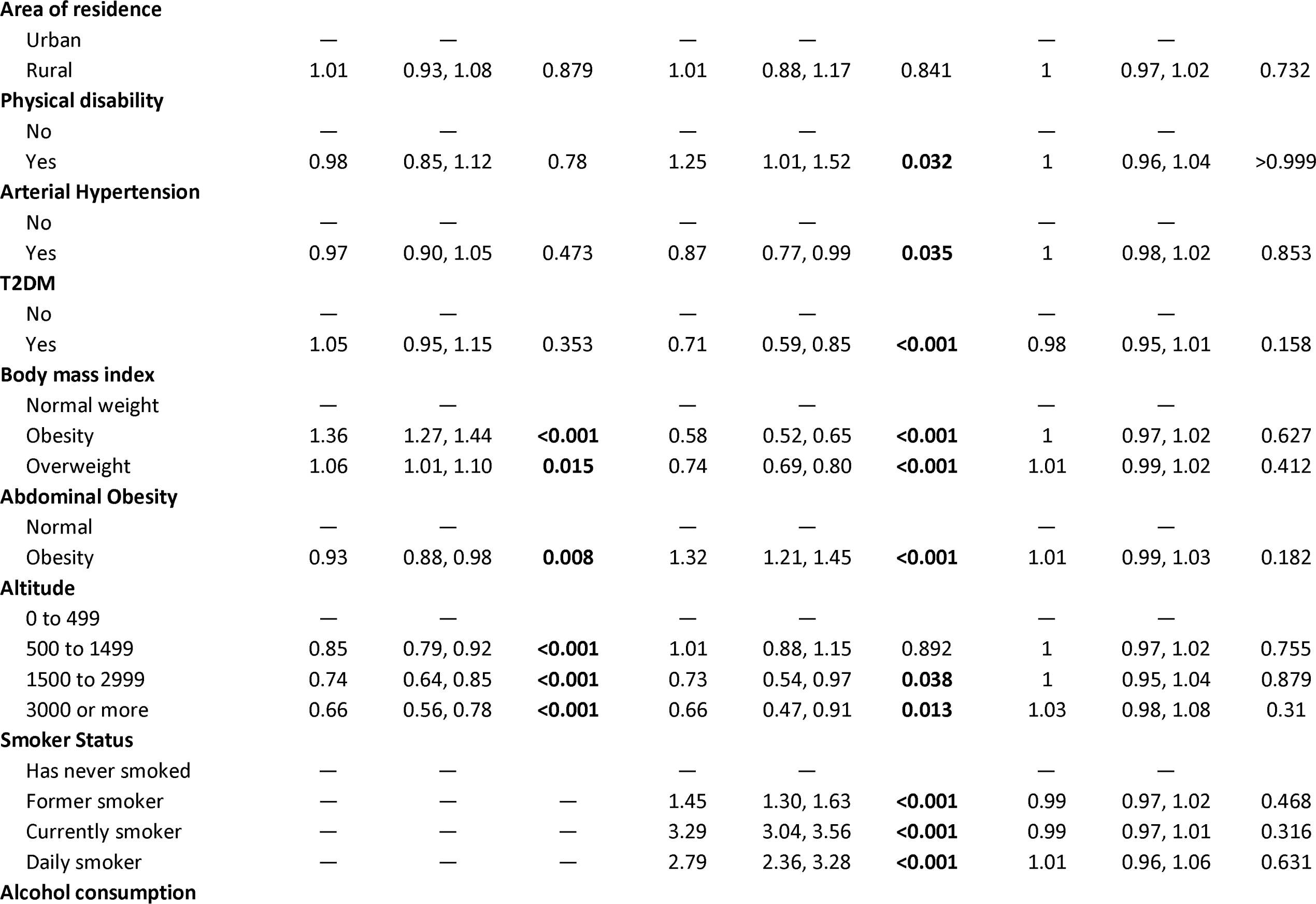
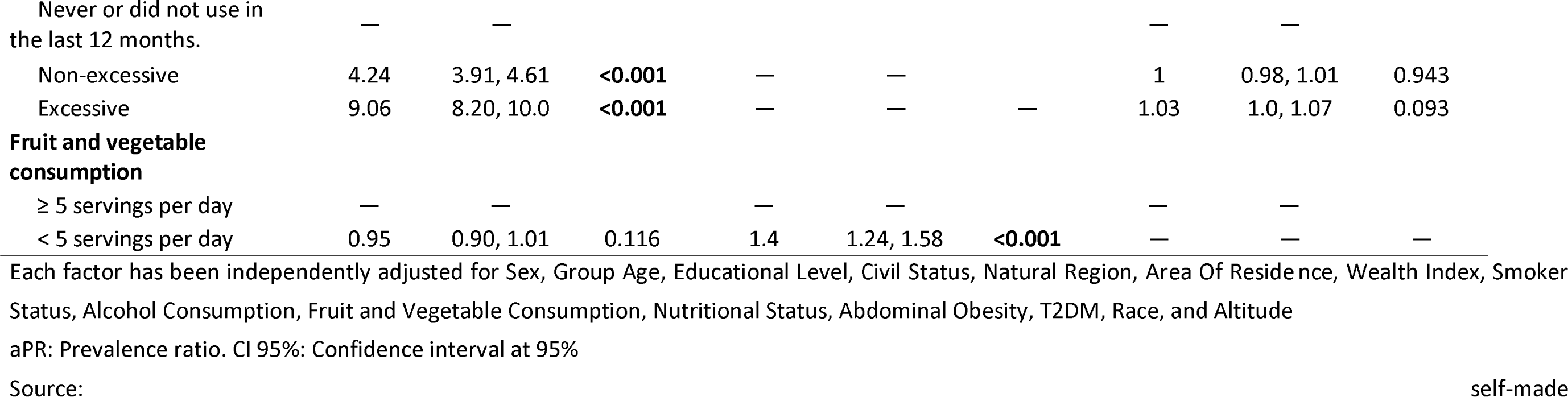
Multivariable regression analysis of factors associated with lifestyles.

**Table 03.**
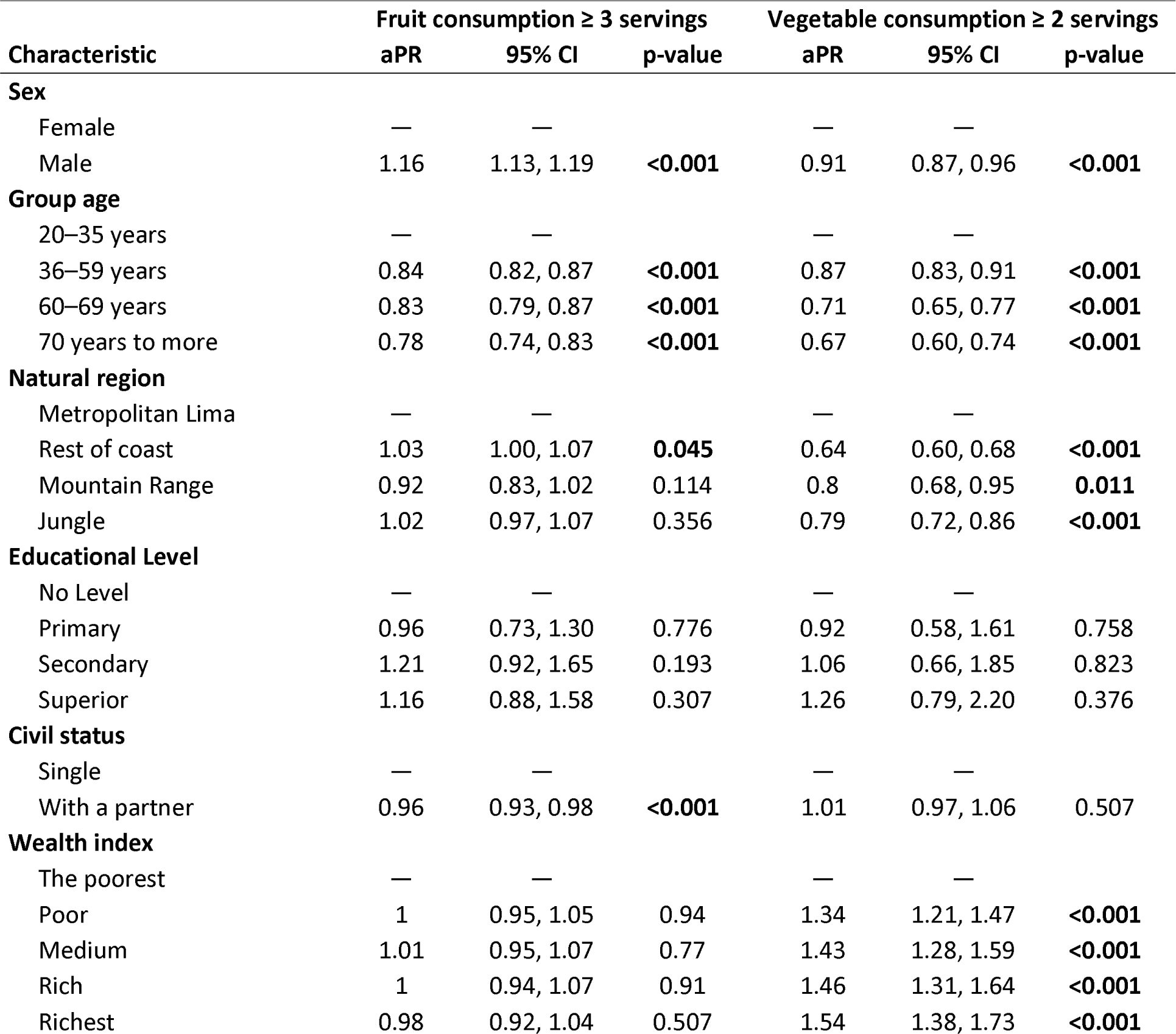
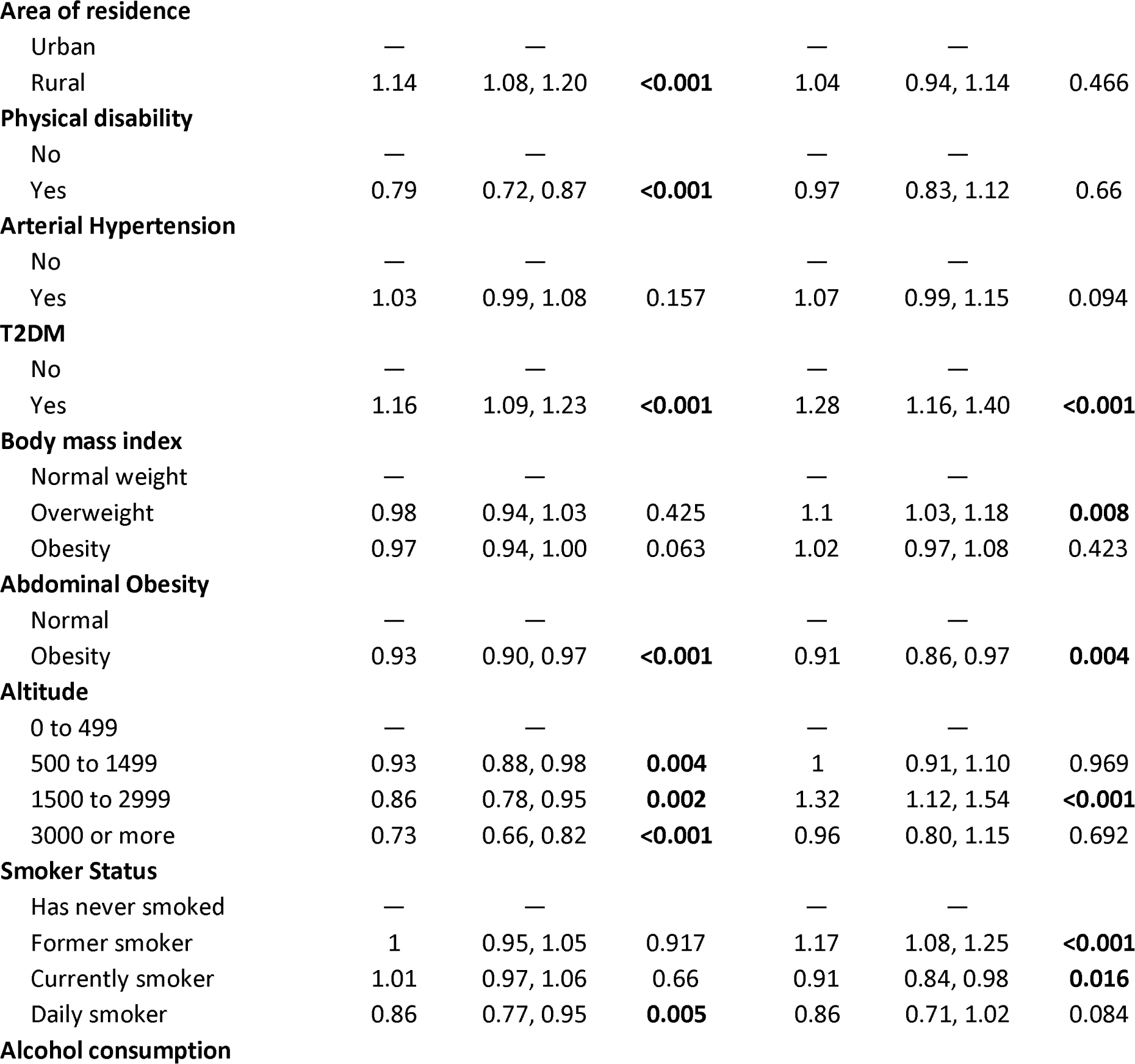
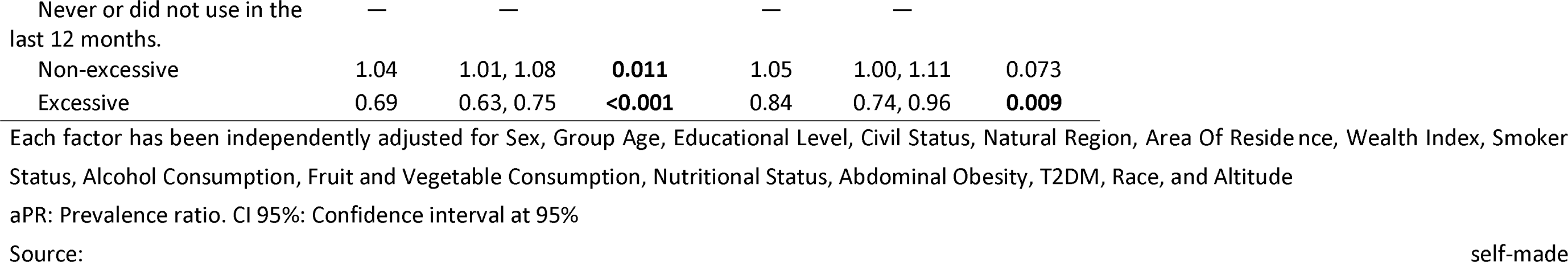
Multivariable regression analysis of the factors associated with the consumption of fruits and vegetables.

Men are more likely to be current or daily smokers compared to women. Age is also a significant factor, with a decrease in the likelihood of smoking as age increases. Living in the jungle is associated with a higher probability of being a smoker than living in Metropolitan Lima.

On the other hand, being male increases the likelihood of both non-excessive and excessive alcohol consumption compared to women. The prevalence of excessive consumption is higher in the wealthiest group compared to the poorest. The presence of arterial hypertension and T2DM is inversely associated with excessive alcohol consumption.

Likewise, significant differences by gender were found in the consumption of fruits and vegetables, with men being less likely to consume the required amount. Older age groups showed a slight tendency towards higher consumption of fruits and vegetables compared to the reference group of 20 to 35 years.

### Factors Associated with Fruit and Vegetable Consumption (Separately)

The regression analysis for fruit consumption (≥ 3 portions) indicates that men are likelier to reach the recommended threshold of at least three portions per day than women. This trend is significant and suggests differences in dietary behavior between genders. Age plays an inverse role in fruit consumption, with a decreasing tendency in the likelihood of consuming the recommended amount as age increases, consistent across all older age groups. Regarding geographic location, residents outside of Metropolitan Lima, especially in the coastal region, show a slight inclination towards higher fruit consumption, though the significance is modest. Rural life is also associated with a higher probability of adequate fruit consumption. However, factors such as educational level and marital status have limited or no significant impact on fruit consumption.

Regarding vegetable consumption (≥ 2 portions), the results reveal an opposite trend in sex, as men are less likely than women to consume at least two daily portions of these. The probability of adequate vegetable consumption significantly decreases with age. Regionally, both the coastal region and the jungle have a substantially lower likelihood of meeting vegetable consumption recommendations than residents of Metropolitan Lima. As the wealth level increases, so does the probability of consuming vegetables. Variables like physical disability and the presence of T2DM had no significant impact on vegetable consumption, while abdominal obesity showed an inverse association with this eating behavior. Finally, altitude plays a role in vegetable consumption, with a notable increase in the likelihood of adequate consumption at intermediate altitudes.

## Discussion

### Prevalence and Trend of Smoking Status

While the Tobacco Atlas indicates that worldwide tobacco consumption prevalence fell from 22.7% in 2007 to 17% in 2021, population expansion has primarily maintained the absolute number of users at a high level, with over three-quarters of male daily smokers residing in nations possessing medium or high human development indices. According to the World Health Organization’s 2020 report, approximately one-fifth of the global population was estimated to have engaged in the use of tobacco during that year, with over one-third of males and just under one-tenth of females reported to have participated in the practice worldwide.

The observed trend in daily and current smoking prevalence suggests an interesting dynamic over the years. While public health policies aimed at curbing smoking and raising risk awareness seem to have gradually impacted daily proportions over time, the ongoing drop also suggests a collective shift is transpiring in recognizing smoking’s health perils. This phenomenon aligns with global trends reported, where smoking prevalence has decreased in most countries due to effective tobacco control policies and changes in social norms ^(3,11)^.

The decrease in daily and current smokers during 2020, followed by an increase in 2021 and 2022, can be explained by various factors related to the COVID-19 pandemic. A systematic study found that the pandemic and governmental policies to manage it have influenced tobacco consumption behaviors. The reduction in 2020 was likely related to fear of COVID-19 infection and restrictions on social gatherings, which might have limited availability and opportunities for smoking ^(12)^. Additionally, alcohol availability in the country is straightforward ^(13)^.

In 2021 and 2022, the increase in smoking prevalence might be associated with the easing of restrictions and a sense of returning to "normality," especially after the introduction of vaccines. Moreover, the cumulative effects of stress and mental health issues related to the pandemic could have led some people to resume or increase their tobacco use as a coping mechanism ^(14)^.

A study conducted in China during the pandemic showed that, after the outbreak nationwide, more individuals quit smoking compared to those who started, and smokers significantly reduced the number of cigarettes consumed. While there existed considerable variances between persons, the research uncovered that gentlemen experienced more difficulties in diminishing their usage of tobacco compared to ladies. This study further observed that a more substantial decrease in tobacco consumption correlated more strongly with higher levels of fulfillment regarding bodily health and emotional wellness ^(14)^.

These findings suggest that public health policies during and after the pandemic should consider differences in individuals’ vulnerability and leverage psychological factors that can facilitate behavioral change. Concern about health risks associated with COVID-19 due to smoking may have been a significant factor in smokers’ behavior change.

### Prevalence and Trend of Alcohol Consumption

Regarding the global prevalence in the country, alcohol consumption in the United States and worldwide presents a diverse picture. The National Survey on Drug Use and Health estimated that approximately four in five individuals aged twelve or older residing in the United States had engaged in the assessed behaviors. A survey found that while the majority of respondents acknowledged having drunk alcohol at some point in their lives, a smaller yet still significant percentage indicated engaging in heavy drinking within the last 30 days, according to the study’s findings. In comparison to the estimated worldwide prevalence of alcohol use disorders at 1.4% with substantial disparities between nations ranging from that figure upwards, countries such as Russia have significantly higher rates, such as 4.7%, as recently quantified ^(16)^. Compared to Peru, the prevalence of alcohol consumption remains, as other studies have pointed out ^(17,18)^. The differences in alcohol consumption prevalence between countries and within the same country between genders highlight the need for culturally adapted and gender-sensitive prevention and treatment policies and programs to address the unique challenges each population faces concerning alcohol. Mitigating the adverse health and social consequences of alcohol consumption necessitates crucial efforts to curb its damaging impacts on individual and community wellness.

On the other hand, the peak in excessive alcohol consumption observed in 2020 could be related to the stress and anxiety generated by the COVID-19 pandemic. A study from Italy using hair analysis to assess alcohol intake reflected changes in drinking behavior during the pandemic, supporting the notion that exceptional circumstances can influence alcohol consumption ^(19)^. Research into coping during the pandemic further revealed that while some people increased alcohol use to handle stress, others lessened or quit consumption owing to worries for well-being and limits on procuring it ^(14)^. The study’s revelations indicate the pandemic could have triggered considerable modifications in alcohol utilization, demonstrating humanity’s intricate reactions to worldwide disruptive phenomena and emphasizing the necessity of contemplating such determinants in public health policy formulation.

### Prevalence and Trend of Fruit and Vegetable Consumption (Together and Separately)

While global fruit and vegetable intake trends have demonstrated considerable fluctuation, consumption levels diverge substantially between nations, with many experiencing notably inadequate amounts.

Fruit and vegetable intake across nations diverges widely as consumption levels differ significantly depending on location. For example, data from the United States from 2015 to 2018 indicates that while a substantial majority of around 95% of adults consumed some vegetables daily, a smaller yet still sizable portion consisting of approximately two-thirds ate some fruit on a given day as well, according to the cited source ^(20)^. A comparison of fruit and vegetable consumption across several nations in 2019 revealed that while Turkey’s intake was 32.87/122.33 kg per person, amounts varied considerably between 19.97/26.97 kg and 53.15/77.13 kg in Brazil, Canada, China, France, Greece, England, and the United States ^(21)^. A systematic review discovered worldwide vegetable consumption tends to be deficient, with many low- and middle-income nations reporting merely 1 to 2 portions daily ^(22)^. A 2009 study that recruited 52 primarily low and middle-income countries found between 77.6% to 78.4% of men and women consumed less than the recommended five daily portions of fruits and vegetables, demonstrating while this work is dated, the current situation likely differs little from what was occurring at that time ^(23)^.

Many factors potentially contribute to why individuals may ingest more servings of fruits instead of vegetables regularly. A commonly cited justification provided by consumers for their lack of fruit and vegetable consumption was found by one study to be the cost of these healthful foods ^(24)^. Another study found that many people do not eat vegetables in their natural low-fat and calorie form, as they would with fruits. French fries are the most popular vegetable option among Americans ^(25)^. Nonetheless, research has found that amongst United States adults over the age of fifty, there has been a noticeable decline in the regular eating of vegetables, which data suggests may contribute to the overall decrease in vegetable consumption nationwide.

While the consumption of fruits tends to surpass that of vegetables across many nations due to an assortment of interconnected causes, the precise rationales behind such dietary variances are likely influenced by a country’s unique cultural environment and the characteristics of its resident groups. There are several reasons behind the disparity in consumption levels between fruits and vegetables. These include financial considerations, individual preferences regarding flavor, and how these plant-based foods are ready and partaken.

### Factors Associated with Smoking Status

Numerous studies have conclusively demonstrated in their analysis of smoking habits that identifying as male correlates more strongly to an increased chance of being an everyday or consistent smoker. Extensive research has explored the widespread occurrence of tobacco consumption and how its correlation with gender and age has differed. Research has disclosed, for example, that gentlemen, as a rule, partake in all sorts of tobacco at a rate surpassing that of ladies. The findings of another study ^(27)^ demonstrated through its results that roughly sixteen percent of adult males and fourteen percent of adult females in the United States were individuals who smoked cigarettes, according to the data collected. It is plausible that the variances stem from interweaving bodily, societal, and conduct-related influences. While neuroimaging research indicates smoking may stimulate the gratification routes in men to a higher degree than in women, consistent with the notion that men smoke more for nicotine’s reinforcing impacts, women are suggested to smoke additional for mood regulation or as a response to cigarette-related prompts. Furthermore, the comparative analysis of seven European nations regarding beliefs about smoking suggested that views diverge about sex, and such perspectives are enmeshed within the overarching social and cultural environment. For example, in countries with more gender-equal cultures, one might expect less pronounced differences in beliefs about smoking between men and women ^(28)^. This variation in cultural context and access to prevention and treatment resources across regions could provide insight into disparities in smoking prevalence between areas, like the likelihood of smoking being higher in the jungle than others due to differing circumstances.

A cross-sectional study of adults in the United States examined how age potentially impacts various factors. Using National Surveys from 2011 to 2022 found that adults under 40 years experienced dramatic decreases in smoking prevalence over the last decade, particularly among those with higher incomes. In contrast with the rapid declines seen in younger age groups, there were comparatively sluggish decreases noticed among the 40 to 64-year-old demographic, with no reduction in smoking habits for those aged 65 or above ^(29)^.

Ultimately, the propensity for more excellent smoking rates corresponding to augmented prosperity may symbolize the intricate interplay of socioeconomic elements linked with tobacco consumption. The research proposes that one’s socioeconomic position in society carries considerable influence, as studies have often found smoking rates to be more prevalent among those holding lower rungs on the socioeconomic ladder. Yet specific investigations have arrived at the antithetical determination. While this pattern may not be entirely consistent, various studies have shown that it appears susceptible to modulation by diverse contextual circumstances and lifestyle influences ^(30,31)^.

Investigations into the interplay between alcohol intake and cigarette smoking habits have revealed, through numerous examinations, a notable correlation between the two practices. Their research, as published in Shiffman and Balabanis ^(32)^, demonstrated that the rate of smoking is substantially greater, coming in 75 percent above, among those who drink in contrast to those who do not imbibe. The study further illuminated a correlation between consumption amounts and co-dependence, whereby the most substantial imbibers exhibited a propensity towards the most significant usage of tobacco products, as was noted in reference thirty-two. Another study published by Jiang et al. ^(33)^ reported that smoking cigarettes is strongly associated with alcohol consumption. The study found that smokers, hefty drinkers, are more likely to smoke than non-drinkers and that the association between tobacco and alcohol use becomes stronger with heavier use of either substance.

A recent time-series analysis of shifting population patterns in England, as featured in Addiction, detected a noteworthy link between tobacco use and heavy alcohol intake to a personal degree. The study assessed whether a similar association is maintained at the population level over time and found positive associations between smoking and drinking parameters ^(34)^. A longitudinal investigation conducted by Hart and colleagues ^(35)^ tracked the intertwined impacts of cigarette consumption and alcohol usage across three decades, determining that nicotine and ethanol’s neurological processes seem to augment one another reciprocally and that the tandem practice of drinking and smoking has profound ubiquitous cultural relevance as an accepted social behavior predominantly within public houses, taverns, and nightclubs globally. A recent study published in Nature found that most genetic associations between drinking and smoking have similar effects across different ancestries, further supporting the strong correlation between the two behaviors ^(36)^.

While the data indicates initiatives to curb cigarette use among younger demographics show promise, more work appears imperative to further lessen tobacco consumption among older age groups, as suggested by these results. Furthermore, interventions should consider differences between sexes and responses to specific treatments, as observed in the case of tobacco-dependence medications. For instance, varenicline has demonstrated greater short- and medium-term efficacy among female smokers, though one-year abstinence rates are comparable between men and women.

### Associated Factors with Alcohol Consumption

Research into gender differences in patterns of alcohol consumption and the epidemiology of related harms has revealed that men demonstrate a propensity for drinking behaviors linked to both moderate and heavy usage at increased levels, a finding consistent with studies showing distinctions between how each sex experiences use and the problems stemming from alcohol. At the same time, alcohol consumption habits in the United States differ between genders; males, on average, drink more regularly and in more significant amounts than females, consuming nearly triple the amount of pure alcohol annually. Unfortunately, statistics demonstrate that males encounter heightened risks of facing arrest for drunk driving, needing emergency medical care for alcohol-caused injuries, and struggling with alcohol use disorders, more so than females. Gender variations in alcohol intake have been extensively recorded internationally, exhibiting dependable uniformity on a global scale. While statistics show males tend to imbibe and overindulge in alcoholic beverages more frequently than females, with a reported as doing so excessively, the reasons behind such gendered differences in drinking habits remain widely debated ^(37)^.

According to the National Survey on Drug Use and Health conducted in the United States, the study found over three-quarters of men and women aged 12 and above, precisely 79.7% of males and 76.9% of females, reported having consumed alcohol at some point in their lives. Furthermore, the report also indicated that in the past month, 5.8% of individuals aged 12 and over had partaken in a substantial level of alcohol consumption, with the percentage of men in this age range reporting excessive alcohol intake, 7.3%, surpassing that of women, 4.4% ^(15)^. While alcohol consumption tends to decrease with age, as excessive drinking proves less common among elderly demographics, this phenomenon lends credence to the proposition that advanced years correlate with more temperate alcohol intake.

Furthermore, research into alcohol abuse patterns among various Peruvian demographic groups discovered that individuals possessing more excellent academic qualifications and belonging to more affluent socioeconomic classes exhibited a more widespread issue with excessive drinking habits, according to the findings reported. Additionally, the research uncovered that the inverse relationship between health issues and excessive alcohol usage is reinforced by the discovery that hypertension and type 2 diabetes demonstrate an opposing relationship with heavy alcohol consumption as well ^(38)^.

There exists an intricate interconnection between alcohol intake and socioeconomic circumstances, as this association encompasses numerous intertwining aspects. Research into the relationship between wealth and alcohol consumption has found evidence that individuals of higher economic status are more prone to engage in immoderate drinking behaviors. For example, one study found that a person’s socioeconomic standing had a direct, positive link to the amount of alcohol they drank, demonstrating that higher-income individuals tended to consume more alcoholic beverages than their lower-earning counterparts, as cited in source thirty-nine. Likewise, Collins S’s ^(40)^ analysis revealed that parental wealth and education were indicative of alcohol consumption in young adults, with those from higher socioeconomic families reporting higher intake and those with greater family affluence disclosing more frequent bouts of excessive drinking.

Regarding obesity and alcohol consumption, related studies have been found, though more on how the latter leads to obesity. A study in the Irish adult population showed that harmful alcohol consumption was associated with both a high BMI and a large WC. Furthermore, the findings noted that individuals who consumed alcohol with greater regularity were more inclined to have a more substantial waist circumference. In contrast, those who frequently partook in alcoholic beverages were less prone to obesity, as determined by their BMI measurement. That same study discovered a notable inverse association between increases in a patient’s BMI and reductions in their alcohol use, suggesting that higher levels of obesity correlate with lower amounts of drinking, according to the reference. While the findings highlighted that heavy drinking notably increased the likelihood of obesity for specific Korean individuals, it is essential to acknowledge the research demonstrating those consuming over 28 beverages weekly faced considerably elevated risks of weight gain. While moderate weekly alcohol consumption showed no correlation, heavier social drinking patterns were linked to an increased likelihood of obesity. In contrast, the light-to-average alcohol consumption group (0 to <28 standard drinks per week) was associated with a lower risk of diabetes ^(43)^.

Nonetheless, these differences found regarding what type of obesity consumes more or less alcohol may be because BMI and WC are indicators that reflect different aspects of obesity. Whereas BMI offers a general metric of body weight accounting for height, WC serves as a more targeted signpost for belly fat levels. Visceral abdominal fat poses grave metabolic and cardiovascular dangers in a manner strongly correlated with its association within the region. Those with a higher waist circumference, signifying more belly fat buildup, could exhibit divergent risk behaviors relative to their peers with a comparable body mass index yet dissimilar overweight allocation, such as elevated alcohol intake ^(44)^.

Studies show that individuals who fail to meet daily recommendations for fruit and vegetable intake are more prone to engage in binge drinking behaviors. The study discovered that in lower-income populations in particular, a tendency existed such that increased intake of fruits and vegetables correlated with decreased alcohol consumption, and vice versa. While this finding indicates the connection between one’s diet and alcohol intake could carry more import where economic alternatives remain limited, it deserves expanded consideration across a broader range of circumstances. Low fruit and vegetable intake has been tentatively linked to greater alcohol use, possibly owing to less health-conscious habits overall. Those failing to incorporate sufficient fruits and vegetables into their diets may lack commitment to nutritional balance and wellness, which could relate to engaging in unhealthy habits like heavier drinking. Socioeconomic hardships could exacerbate matters as those with fewer means may face more significant challenges in obtaining nutritious sustenance while perhaps being more prone to using alcohol as a strategy for dealing with difficulties or finding diversion. This interconnection between diet and alcohol consumption habits underscores the complexity of health behaviors and how they can influence each other.

### Associated Factors with Fruit and Vegetable Consumption

Regarding eating fruits and vegetables, age plays a notable role in predicting amounts, as there is a propensity for greater intake among more advanced age brackets. While fruit and vegetable intake varied little between demographic groups, with no substantial divergences seen regarding gender, education, marriage, income, or place of living, deeper analyses may yet illuminate meaningful distinctions within these categories.

The connection between one’s age and how much fruit and vegetables one consumes has been explored in different settings, with tendencies noted for increased intake the older individuals become. That British study examined how intake of fruits and vegetables might vary throughout childhood, tracking any alterations from youth to the later high school years. They found that consumption patterns change and can be influenced by several factors throughout development, such as food neophobia and increased peer interaction ^(45)^.

Additionally, a Canadian study identified that older individuals are more likely to consume fruits and vegetables than other age groups. This study also highlighted the association between higher fruit and vegetable consumption and certain healthy lifestyle behaviors, including educational level and marital status ^(46)^.

These findings suggest that age may be a significant predictive factor in fruit and vegetable consumption, with a tendency for increased consumption in older age groups. This could be due to greater health awareness in older ages, changes in food tastes and preferences, or even differences in dietary habits and lifestyle throughout a lifetime.

### Fruit and Vegetable Consumption Patterns

While current research has found that males tend to eat fewer vegetables than females, claims proposing greater male consumption of fruits over females contradict what studies have revealed. However, research carried out in Finland and the Baltic nations, plus a pan-European investigation, has shown that females consume more fruits and greens than males on average. There is a possibility that the variances seen in consumption habits between groups stem from the social standards and customary ideals concerning gender roles that different cultures espouse concerning sustenance and well-being. While females tend to focus more intently on nutritional values when considering their food choices, classifying options based mainly on health content, males have been shown to prioritize these concerns less frequently in their decision-making, per the cited studies ^(47,48)^. That same study discovered males in Finland and the surrounding Baltic nations tended to eat more meat, potatoes, bread, and alcohol in their diet compared to females, opting for less fruit, vegetables, fish, poultry, cheese, and sweets than their female counterparts ^(47)^. While research into the connection between fruit and vegetable intake and age has not consistently upheld the notion that advancing years necessarily diminish consumption of these items, specific investigations have found a potential inverse relationship between growing older and the likelihood of eating such healthful foods. The study investigating the relationship between vegetable and fruit intake and age-related cognitive functioning found that while high vegetable consumption may be connected to a slower pace of cognitive decline in older adults, the same connection could not necessarily be made regarding fruit intake alone ^(49)^.

While research into geographic differences in fruit and vegetable intake failed to uncover any studies directly confirming expectations of lower vegetable consumption in coastal, highland, and jungle areas, no evidence was found either refuting the possibility of variation in produce selection between regions. While local availability and differing access to various foodstuffs could impact typical consumption habits, other considerations may also influence patterns.

While specific investigations have revealed a gradient association linking educational attainment, commonly aligned with the financial station, to the intake of fruits and greens, some analyses have demonstrated a connection between affluence and vegetable consumption. Nonetheless, the degree to which this association holds fluctuates between different localities and settings rather than maintaining a uniform linkage everywhere without exception. In some areas, especially those with high availability and consumption of these foods, it has been observed that less educated people consume more fruits and vegetables compared to more educated individuals ^(48)^.

Concerning people with diabetes, specific research on their fruit and vegetable consumption patterns was not addressed in the reviewed studies. While a plant-based menu aligned with standard guidance for those with diabetes may seem the logical choice for some, others still struggle with consistently nourishing their body in a manner befitting their state of health.

Finally, regarding former smokers and their vegetable consumption, no studies directly address this relationship. While ex-smokers potentially embrace healthier habits post-cessation, like boosted intake of vegetables, firm conclusions remain elusive, demanding further exploration to substantiate the notion that quitting smoking may correlate with adopting an overall wellness-oriented lifestyle.

While the study findings showed some correlation between fruit and vegetable intake, it did not definitively link those who partake in one to participate in the other consistently. This finding suggests the existence of differentiated dietary behavior patterns within the Peruvian population. While one who favors the consumption of fruits could logically be viewed as likewise inclined towards partaking of vegetables, as both comprise essential elements of nutritious and well-balanced nourishment, such presumptions remain an oversimplification. Yet this situation prompts crucial interrogations regarding societal tendencies, accessibility, and subjective and communal perspectives on well-being and nourishment. While variations in flavor, feel, and convenience of preparation set fruits and vegetables apart, these dissimilarities of taste, texture, and effort required for cooking could account for the observed phenomenon. Fruits are generally sweeter, more accessible to consume without preparation, and may be more appealing as snacks or meal accompaniments. However, vegetables commonly demand more work before eating and are seen as less enjoyable regarding flavor, notably when placed next to sweeter fruits. Another critical aspect is availability and cost. In some regions, it may be easier or more economical to access certain fruits than vegetables, or vice versa, thus influencing consumption patterns. While cultural traditions and local eating habits undoubtedly affect typical diets in various regions, they may also substantially impact the extent to which specific food categories are commonly consumed.

This finding highlights the need for differentiated approaches in health promotion strategies. It cannot be assumed that effective interventions to increase fruit consumption will be equally effective for vegetables. More personalized and evidence-based public health strategies are required, considering specific preferences and barriers for each type of food. While additional investigation is certainly warranted to gain deeper insight into the underlying motivations behind these eating habits and to craft more impactful initiatives encouraging a more balanced, nutritious way of consuming food

### Public Health Significance of the Findings

The findings of this study, using ENDES data from Peru, have significant implications for public health. By investigating the interconnections between tobacco and alcohol use, fruit and vegetable intake, and diverse sociodemographic and medical characteristics, this wide-ranging analysis provides insightful consideration into how such determinants reciprocally act upon and impact one another within the population of Peru.

The observed decrease in the prevalence of smokers, especially during 2020, suggests a potentially positive shift in public health behaviors. It is plausible that this phenomenon mirrors the effect of policies aimed at curbing tobacco usage and a rising consciousness concerning the health dangers linked to smoking. However, the possibility that the rate of decline fails to meet expectations could suggest that existing tobacco control public health policies may not be wholly successful or universal in their implementation across all demographics.

The increase in the prevalence of excessive alcohol consumption in 2020, followed by a decrease in 2022, could be associated with the COVID-19 pandemic and its psychosocial effects, such as stress and anxiety. This pattern reflects how disruptive global events can influence alcohol consumption behavior and underscores the need for public health strategies that address these factors.

While fruit and vegetable intake remains deficient, possibly implying meaningful obstacles restrict access to these nutritious options, alternative perspectives recognize low consumption may result from other influences yet to be uncovered. Potential reasons include expense, limited accessibility, insufficient understanding of their dietary significance, or customary preferences. Globally, the absence of fruit and vegetable intake represents a pressing issue as kindred patterns have emerged worldwide.

These findings underline the need to review and possibly strengthen current public health policies and strategies. It is crucial to address barriers to adequate consumption of fruits and vegetables and improve the efficacy of tobacco control measures. This may include education and awareness campaigns, ensuring greater accessibility and affordability of fruits and vegetables, and stricter and more widely applied policies for tobacco control. Efforts to improve public health and reduce the burden of diet-related diseases and smoking are indispensable, as through such initiatives, we are better positioned to foster healthier communities and enhance the quality of life for citizens.

### Study Limitations

While this cross-sectional observational study may discern associations, firm conclusions regarding causality remain elusive given the inherent constraints of its non-experimental design. It cannot be determined whether the identified factors associated with health behaviors are causative of these behaviors. While data on tobacco, alcohol, and fruit/vegetable intake rely on self-reported recollections that may be imperfect due to faulty memory or a tendency to provide socially acceptable answers, this methodology is also prone to distort consumption levels through overestimation or underestimation. While definitions of excessive alcohol use, fruit and vegetable intake, and smoking status are subject to individual interpretation, standardizing these characterizations could help ensure uniformity in how such lifestyle factors are assessed. In addition, the lack of specific details on amounts and frequencies may limit the accuracy of these measures. While the study examines various related attributes, there exists a potential that other undetermined or unattributed confounding elements could impact the outcomes. While the findings concerning Peru offer valuable insights, one must be cautious not to overextend the implications of these results to other dissimilar settings or groups without further study. Cultural, socioeconomic, and health policy variances across settings may impact the degree to which resultant inferences can be extrapolated beyond the present circumstances. Throughout the extended timeframe of the research project, which encompasses multiple years from 2014 through 2022, the observed behavioral tendencies may have evolved over the progression of events due to shifting external circumstances such as modified health policies, fluctuating social inclinations, or worldwide occurrences, for example, the global pandemic.

### Conclusions

The current study observed slight changes in the prevalence of daily and current smokers. Additionally, an increase in excessive alcohol consumption in 2020, followed by a decrease in 2022, was found. The study draws attention to the troubling scarcity of fruit and vegetable intake within the population, underscoring potential health risks posed by inadequate consumption of these nutritious foods. Given the importance of these in maintaining a balanced and healthy diet, there are potentially significant ramifications for public health should their consumption decline substantially. The study found substantial divergences in wellness practices relating to gender, age, and area but also highlighted how accounting for such elements in public health intervention strategies is paramount by emphasizing the need to consider those variances.

While decreases in smoking rates have been observed, maintaining and intensifying tobacco regulation remains paramount, especially for demographics where prevalence persists at unacceptably high levels or declines less than desired. Furthermore, developing and implementing public health strategies necessitates specifically addressing the observed changes in alcohol consumption patterns, especially relating to the pandemic and its aftermath, with nuanced consideration. In addition, implementing programs and campaigns raising awareness of the importance of fruit and vegetable consumption could help overcome obstacles like expense, access, and customary tastes. Finally, further studies should be encouraged to understand the observed trends better and continuously monitor the effectiveness of public health interventions.

## Acknowledgments

A special thanks to the members of Instituto de Investigación en Ciencias Biomédicas de la Universidad Ricardo Palma, who provided valuable comments during the preparation of this study.

## Informed consent

Informed consent was obtained.

## Data availability

The data supporting the findings of this study can be accessed by the original research paper at the following link: https://proyectos.inei.gob.pe/microdatos/

## Authors’ contribution

**Víctor Juan Vera-Ponce:** Conceptualization, Investigation, Methodology, Resources, Writing - Original Draft, Writing - Review & Editing

**Fiorella E. Zuzunaga-Montoya**: Investigation, Project administration, Writing - Original Draft, Writing - Review & Editing

**Luisa Erika Milagros Vásquez-Romero**: Investigation, Resources, Writing - Original Draft, Writing - Review & Editing

**Joan A. Loayza-Castro**: Software, Data Curation, Formal analysis, Writing - Review & Editing

**Carmen Inés Gutierrez De Carrillo:** Validation, Visualization, Writing - Original Draft, Writing - Review & Editing

**Stella M. Chenet:** Methodology, Supervision, Funding acquisition, Writing - Review & Editing

---

**Supplementary material 01.**
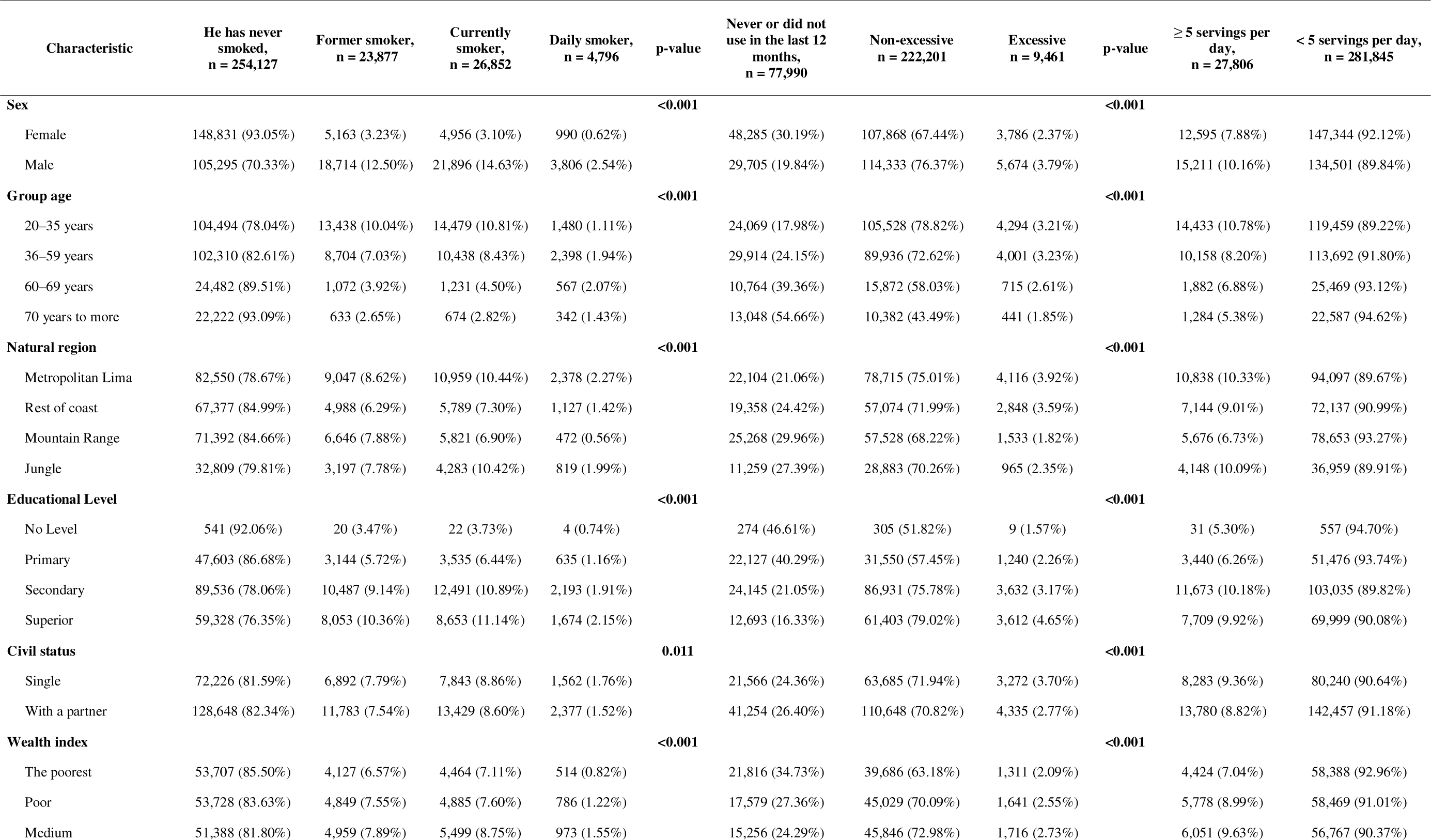
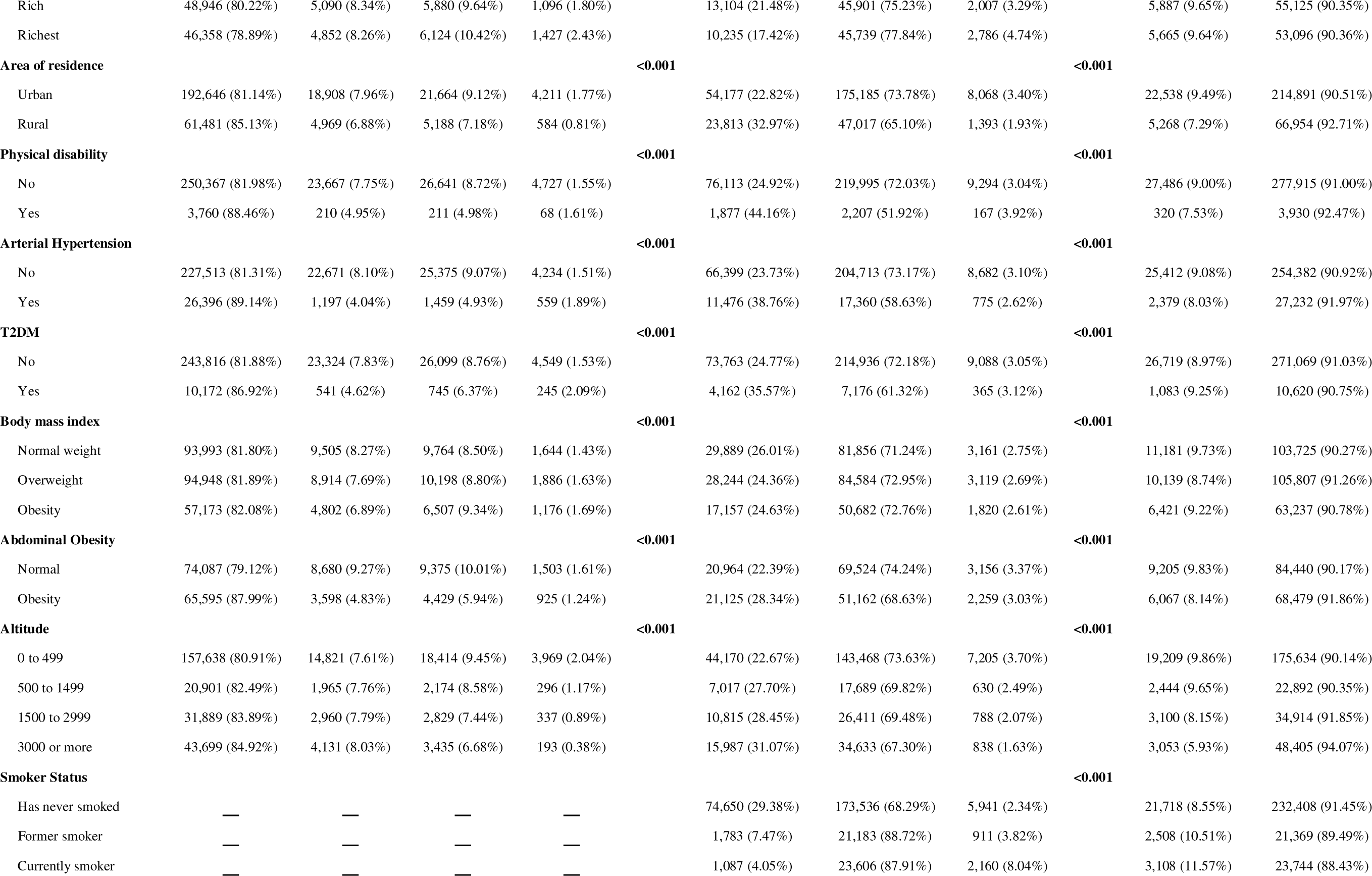
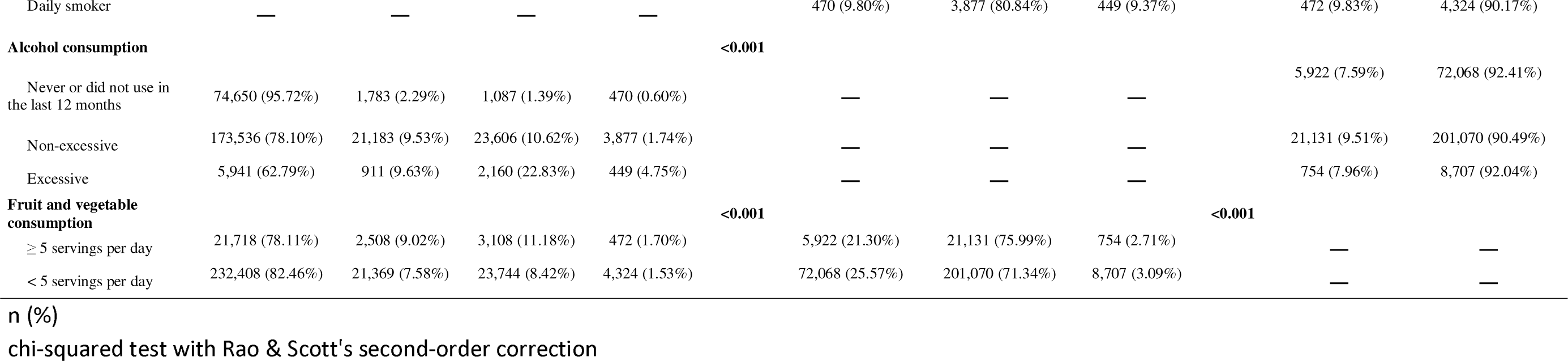
Bivariate analysis of factors associated with lifestyles in the Peruvian population.

**Supplementary material 2.**
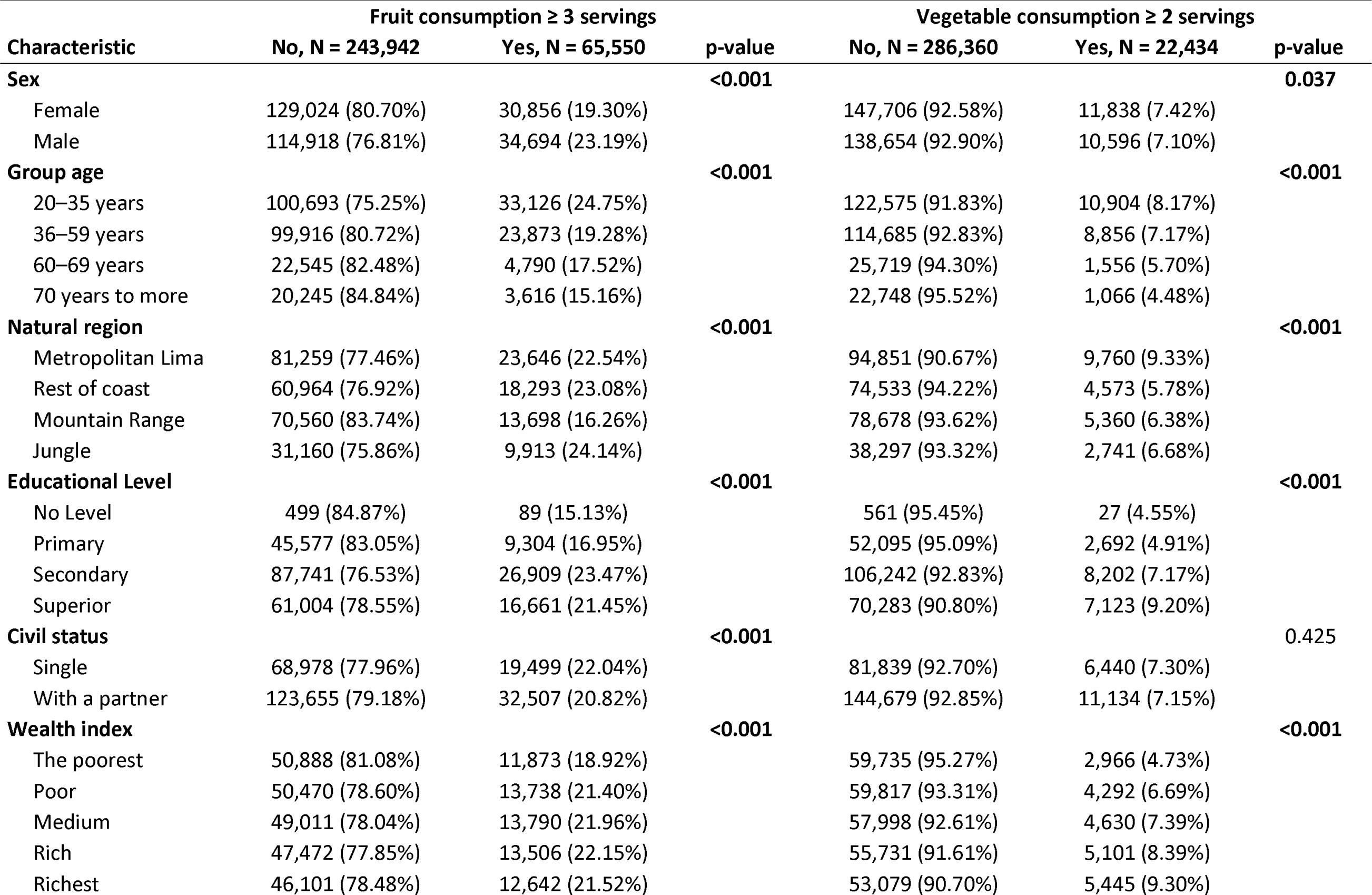
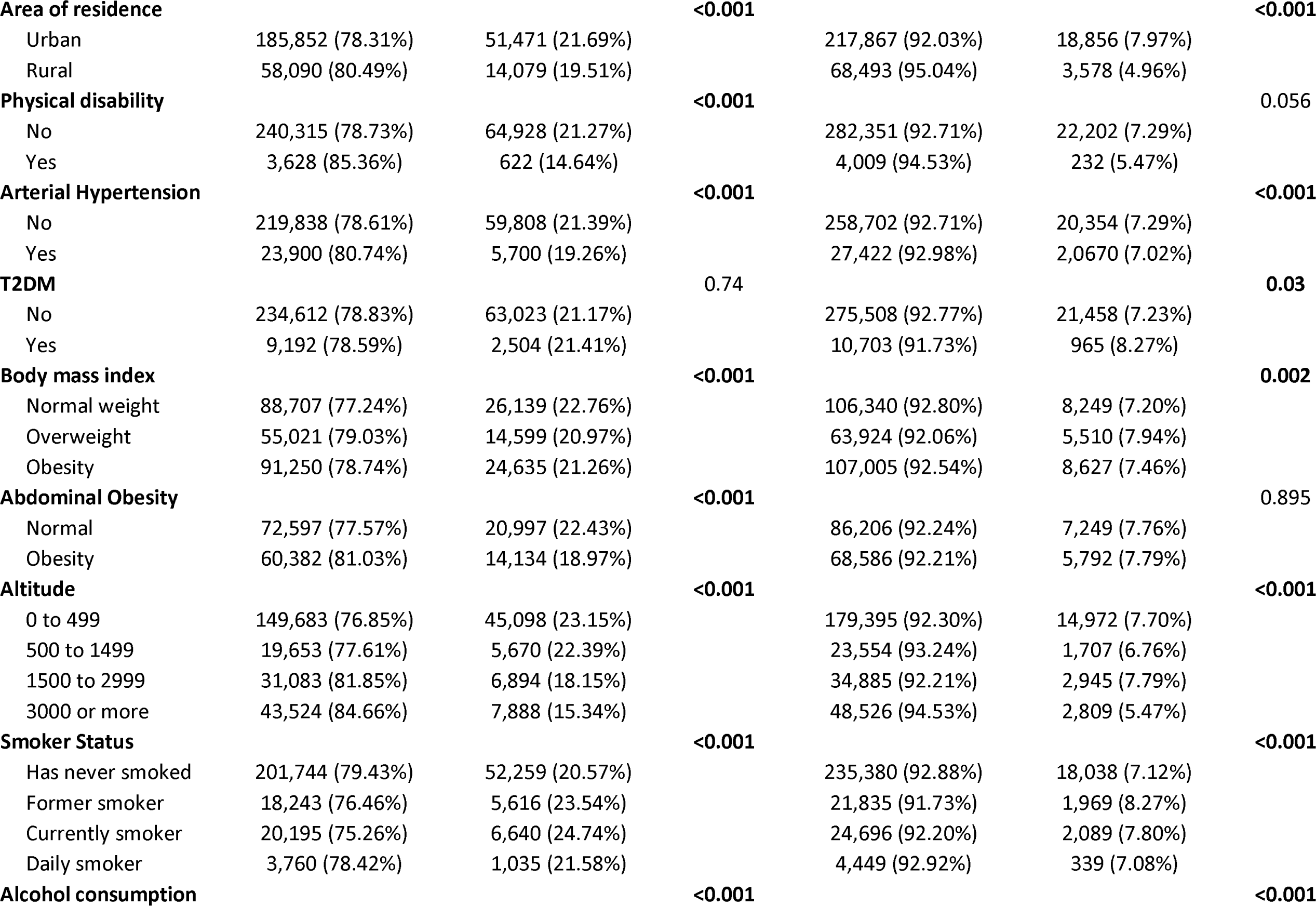
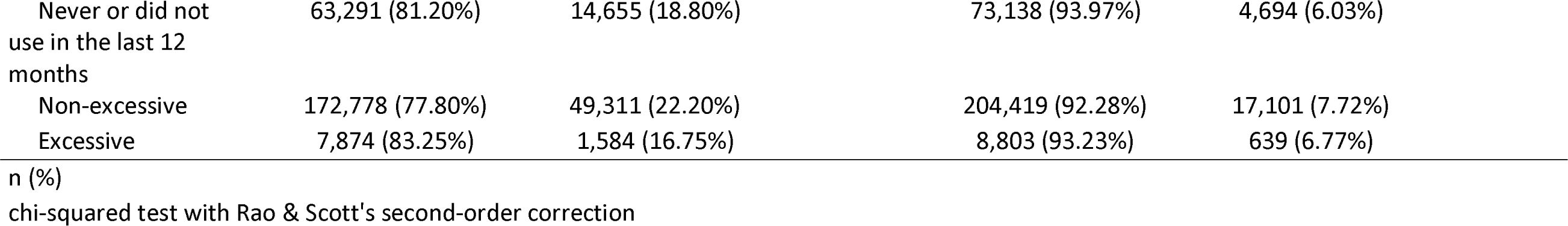
Bivariate analysis of the factors associated with the consumption of fruits and vegetables in the Peruvian population.

